# Hidden in Plain Sight: Epidemiological Signals in Routine Laboratory Data

**DOI:** 10.64898/2026.02.05.26345657

**Authors:** Till Hoffmann, Douaa Mugahid, Jason Olejarz, Angela Neale, Alex Zapf, Ross Molinaro, Marc Lipsitch, Rifat Atun, Yonatan Grad, Sarah Fortune, Rangarajan Sampath, Jukka-Pekka Onnela

## Abstract

Public health monitoring traditionally relies on active reporting from diverse data sources, including clinical and administrative data, disease registries, and population-based surveys. Yet these surveillance methods often face challenges such as incomplete reporting, time lags, and variable population coverage. Meanwhile, diagnostic laboratories routinely generate vast volumes of operational data that are currently untapped for public health monitoring. As these data are not collected for scientific inquiry or population-level surveillance, they often lack formal validation and may contain sensitive information. We developed a Bayesian hierarchical model to decompose aggregated laboratory assay volume data for 1.1 billion clinician-ordered assays across the U.S. from October 2019 to March 2023 into interpretable epidemiological and health system signals. The signals generated by these models were compared with known perturbances to health systems, such as the COVID-19 pandemic. The method does not rely on assay outcomes or individual-level data, providing quantitative signals of epidemiological trends and health system responses while protecting both the privacy of patients and commercially sensitive information. Temporal analysis reveals qualitatively different responses of assay volumes to major public health events, identifying assays whose use paralleled surges in hospitalization rates during the COVID-19 pandemic documented through traditional public health reporting structures. This framework suggests that routine operational data can be used to augment traditional surveillance by identifying anomalous patterns for expert epidemiological investigation. To be truly effective, data from multiple vendors must be integrated to create a comprehensive real-time national or supranational public health surveillance platform.

## Main Text

The foundation of modern public health lies in the ability to rapidly collect, integrate, and analyze diverse data streams with advanced analytic approaches, enabling timely surveillance and informed, targeted interventions tailored to specific populations or health threats. Traditional public health surveillance requires costly acquisition of primary data or painstaking aggregation of retrospective health data across fragmented systems. Meanwhile, diagnostic instruments continuously generate detailed testing records at an unprecedented scale, often millions per day (Mugahid, et al. 2023). These digital data are collected routinely and available in real-time, but access is often limited to private networks. They are rarely used beyond operational monitoring, and their potential for public health surveillance remains untapped. While generated for operational purposes rather than for surveillance—much like search engine queries for disease symptoms (Ginsberg, et al. 2009) or mobile phone data for mobility monitoring (Buckee, et al. 2020)—laboratory data provide a direct link to clinical activity. They are driven by clinicians ordering diagnostic tests based on the presentation of their patients, which, in aggregate, signal health system responses to epidemiological shifts.

To develop a proof of concept for public health monitoring using diagnostic instrument data, we harnessed data from Siemens Healthineers’ Atellica high-throughput platform for automated clinical chemistries and immunoassays. Data were collected in 45 U.S. states between October 2019 and March 2023. Clinical chemistries measure concentrations of substances such as electrolytes, enzymes, and metabolites in blood and other body fluids. Immunoassays detect and quantify specific proteins, hormones, or small molecules like drugs using antibodies. These assays collectively represent a foundational component of the diagnostic work run in hospitals on a routine basis.

The data spanned the COVID-19 pandemic, allowing us to validate our data-driven approach using the events of the pandemic as known public health disruptions. We used assay volume data, i.e., the number of runs of a particular assay. This data was collected as part of routine operations and sent to a backend system for monitoring instruments, resulting in no additional data collection cost for our analysis. However, leveraging these data for public health surveillance necessitates developing reliable data pipelines to transform operational data into consistent, quality-assured datasets. These datasets must be suitable for further statistical analysis to disentangle epidemiological signals from noise and confounders, such as the effect of holidays, instrument maintenance, and specialization of laboratories, e.g., renal laboratories. We focused on aggregated test volume data instead of individual-level outcomes to avoid patient privacy concerns while still capturing data associated with diagnostic needs.

We used data from 97 assay types run on 595 instruments, totaling approximately 1.1 billion assay runs. We aggregated data weekly to reduce the dataset size to approximately 3.2 million records. The aggregation also obviates the need for modeling intra-week variability, e.g., weekday vs. weekend variability in testing. The data are heavy-tailed, noisy, and heterogeneous, necessitating careful modeling to disentangle epidemiological signals from noise.

To decompose these complex data into interpretable factors, we developed a model that could simultaneously isolate the temporal signal of subtle changes in assay use that reflect changing clinical needs, while controlling for the idiosyncratic use of hundreds of instruments, the typical use of different assays, and external factors like holidays. A hierarchical Bayesian model is uniquely suited for this task, as it allows us to model multiple sources of variation in a unified and interpretable framework. The order-three tensor *y*_*ijt*_ represents the number of runs of assay *j* performed by instrument *i* in week *t*, and we used the log-linear model

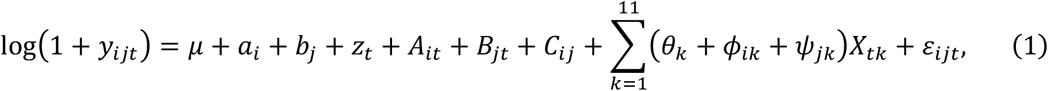

where the log(1 + ·) transform attenuates the effect of outliers while supporting data points with *y* = 0.

Each term represents a different aspect of the data. The term of primary interest for public health monitoring is *B*_*jt*_—the temporal variation in assay volume after controlling for other effects. The parameter μ captures the overall test volume, *a*_*i*_ is an instrument-specific intercept, distinguishing between high- and low-throughput instruments, and *b*_*j*_ is an assay-specific intercept, distinguishing between common assays (such as routine diagnostics) and rarer assays (such as assays for acute conditions). Common temporal effects that affect all instruments and assays are captured by *z*_*t*_, such as seasonal variation. The *A*_*it*_ term captures instrument-specific temporal effects, such as downtime for maintenance. The interaction term *C*_*ij*_ represents specialization of instruments to specific assays. We included fixed effects for federal holidays in the U.S. because we expected laboratory use to be affected by holidays a priori. Specifically, the features *X*_*tk*_ encode federal holidays in the U.S., i.e., *X*_*tk*_ = 1 if week *t* includes holiday *k* and *X*_*tk*_ = 0 otherwise. The coefficients θ_*k*_, ϕ_*ik*_, *ψ*_*jk*_ capture the common, instrument-, and assay-specific responses to holidays, respectively. Residual variation not accounted for by the components is captured by the noise term ε_*ijt*_∼Normal(0, κ_*ij*_) with variance 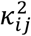 specific to instrument *i* and assay *j*. We chose Gaussian process priors with Matérn-3/2 covariance kernel (Rasmussen and Williams 2006) for the temporal effects *z, A*, and *B* because they naturally model the smooth yet complex dynamics of test utilization. The marginal scale and correlation length are denoted by *σ*^({*z,A,B*})^ and *ℓ*^({*z,A,B*})^, respectively. Hierarchical shrinkage priors were used for the intercept terms *a*∼Normal (0, *σ*^(*a*)^) and *b*∼Normal (0, *σ*^(*b*)^). We fitted the model using variational inference because of the large dataset size and the model having almost 250 thousand parameters (Ranganath et al., 2018).

To analyze the underlying factors driving the use of assays, each additive component of the log-linear model in eq. (1) can be interpreted individually. For example, exp(*z*_*t*_) represents a multiplicative factor scaling the predicted volume in week *t*, controlling for all other factors. A value of 1.5 corresponds to a 50% increase over typical use.

Raw volume data for two assays and two instruments are shown in Figure 1 together with the reconstruction based on components of our tensor decomposition model. Each component represents one aspect of the data, and the scale parameters *σ* can be used to assess the relative contributions of each component (Gelman 2005). Instrument-specific components capture most of the variation because volumes for different assays run by the same instrument are often heavily correlated. The posterior median of *σ*^(*A*)^ averaged across instruments is 0.9155 with 95% credible interval (CI) 0.9138–0.9158. In contrast, volumes for the same assay run by different instruments are only weakly correlated due to common epidemiological drivers. The posterior median of *σ*^(*B*)^ averaged across assays is 0.0504 with 95% CI 0.0502–0.0505. We chose to visualize high-volume instruments in Figure 1 to show data across the entire observation period. The reconstructions are slight underestimates of the data because, as expected, the priors of the Bayesian hierarchical models regularize the instrument-specific random effects *a* of these high-volume systems. As discussed below and shown in panel (b) of Figure 2, assay-specific temporal effects *B* of the two assays shown, C-reactive protein and lipase, were strongly correlated and anti-correlated with weekly national COVID-19 hospitalizations, respectively.

**Figure 1.**
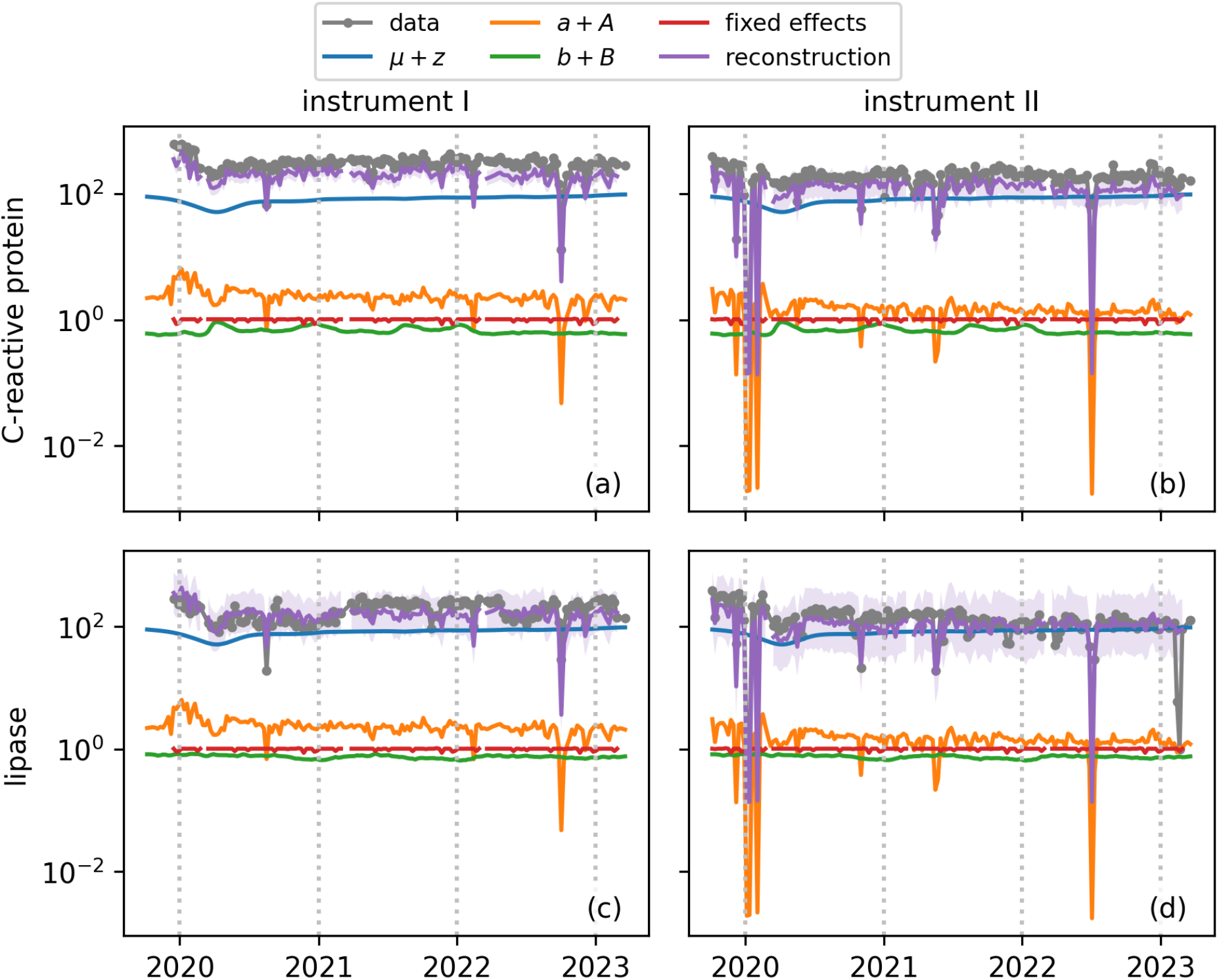
Variational tensor decomposition can reconstruct complex, noisy data for two instruments and assays. Each panel displays the weekly volume of two assays (rows) performed by two instruments (columns) in gray with markers. The common effect μ + z is shown in blue, instrument-specific effects a + A in orange, assay-specific effects b + B in green, fixed effects due to holidays in red, and predictions in purple. The error band corresponds to the 95% credible interval of the posterior predictive distribution.

**Figure 2.**
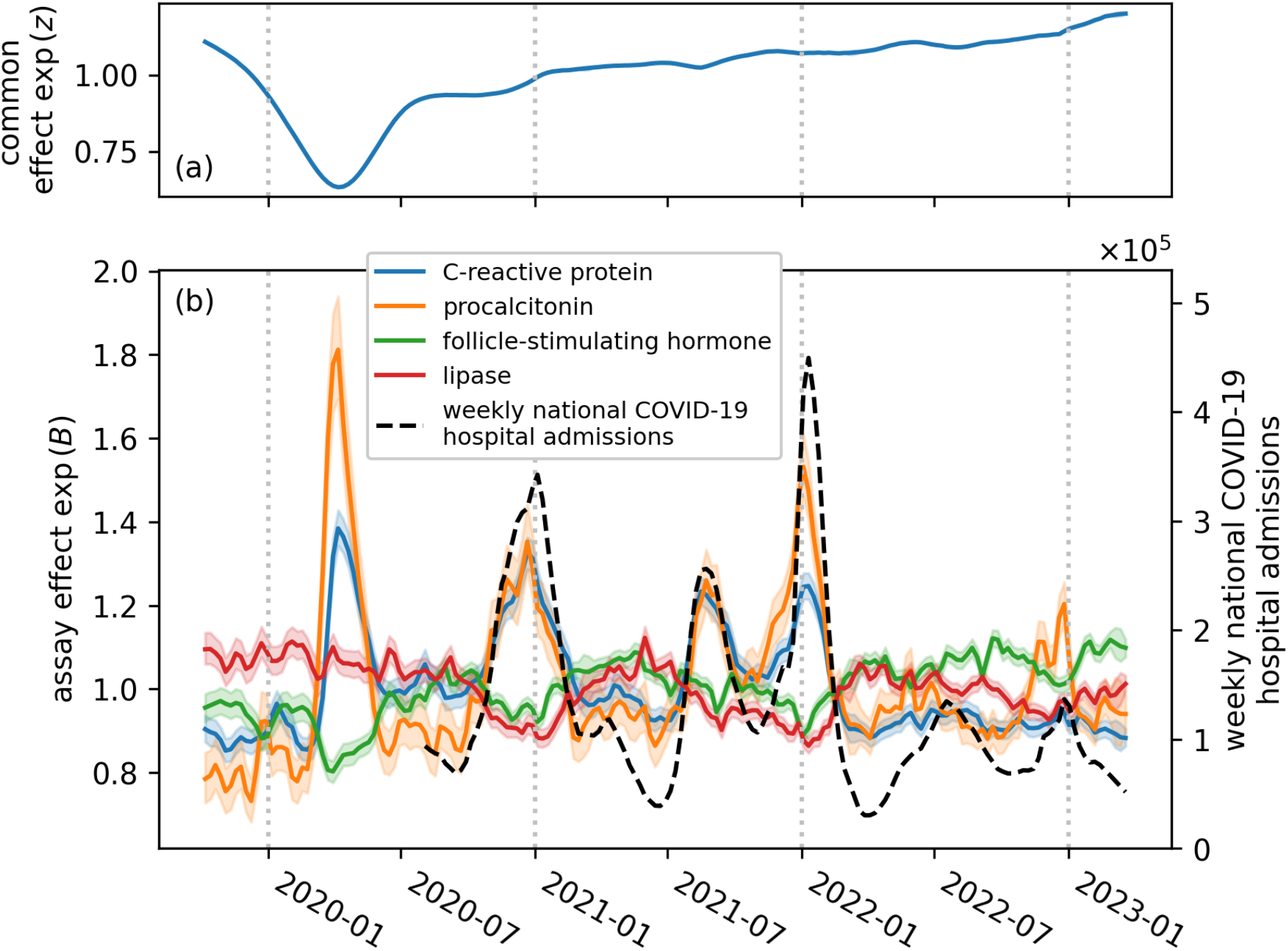
Routine diagnostics and critical care assays react differently to external shocks. Panel (a) shows the posterior median for exp(z) which modulates the assay volume across all instruments and assays and shows a significant reduction during the first COVID-19 wave. Panel (b) shows assay-specific effects as solid, colored lines with 95% credible bands. Weekly national hospital admissions associated with COVID-19 are shown as a dashed black line. On the one hand, routine assays for monitoring reproductive endocrinology and pancreatic disease are suppressed during COVID-19 waves. On the other hand, assays used in critical care management, such as procalcitonin as a sepsis marker, closely track waves. Hospitalization data were not available in early 2020, but the first COVID-19 wave is captured by the signal inferred from operational data. In both panels, a value of 1.0 represents the typical assay volume. A value of 1.5 signifies a 50% increase over the baseline, while a value of 0.75 signifies a 25% decrease.

Careful interpretation of temporal variation in assay use is essential, even for assessing and refining data quality. For example, the analysis revealed a significant, unexplained reduction in assay volumes in the middle of March each year, as shown in panel (b) of Figure 3. After investigation of the data pipeline, we could attribute the reduction to a desynchronization between the backend and instruments after changing to daylight saving time. This issue was not readily discernible from the raw data because the reduction in test volume was obscured by similar effects due to federal holidays. Because the data missingness is not related to the epidemiological situation, we were able to discard the affected data without affecting the analysis (Gelman, Carlin, et al. 2013, ch. 8).

**Figure 3.**
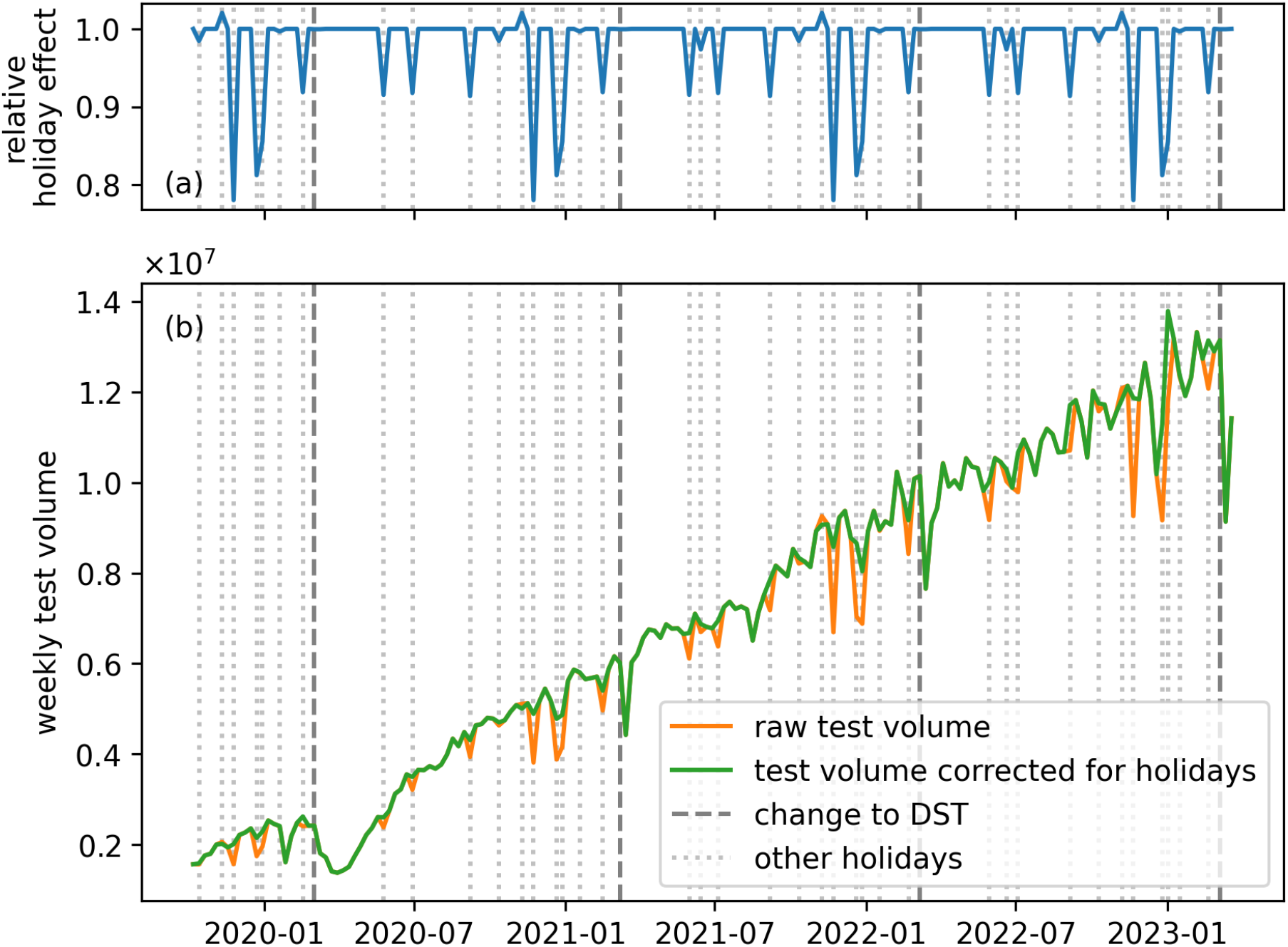
Controlling for holidays reveals missing data due to desynchronization of instruments and backend after change to Daylight Saving Time (DST). Panel (a) shows the inferred relative effect of federal holidays indicated by dotted lines. E.g., during the weeks of Christmas and New Year’s Day, testing volume is estimated to be reduced to approximately 70% of its typical volume. Dashed lines represent the change to DST. Panel (b) shows the raw test volume data and test volumes corrected for the effect of federal holidays, i.e., raw test volumes divided by the relative holiday effects in panel (a). The holiday-corrected time series revealed significant drops in test volume after change to DST which could be attributed to a desynchronization of instruments and backend, resulting in data loss.

To analyze assay-specific effects holistically, we applied a hierarchical clustering algorithm to assays, treating the mean absolute difference between rows of *B* as a similarity metric. The clustering, represented as a dendrogram in Figure 4, reveals both epidemiological trends and linkage inherent to how assays are ordered. For example, assays within different panels—such as the basic metabolic panel, comprehensive metabolic panel, and drug and toxicology panel— cluster together because these assays are typically ordered as a set rather than individually.

**Figure 4.**
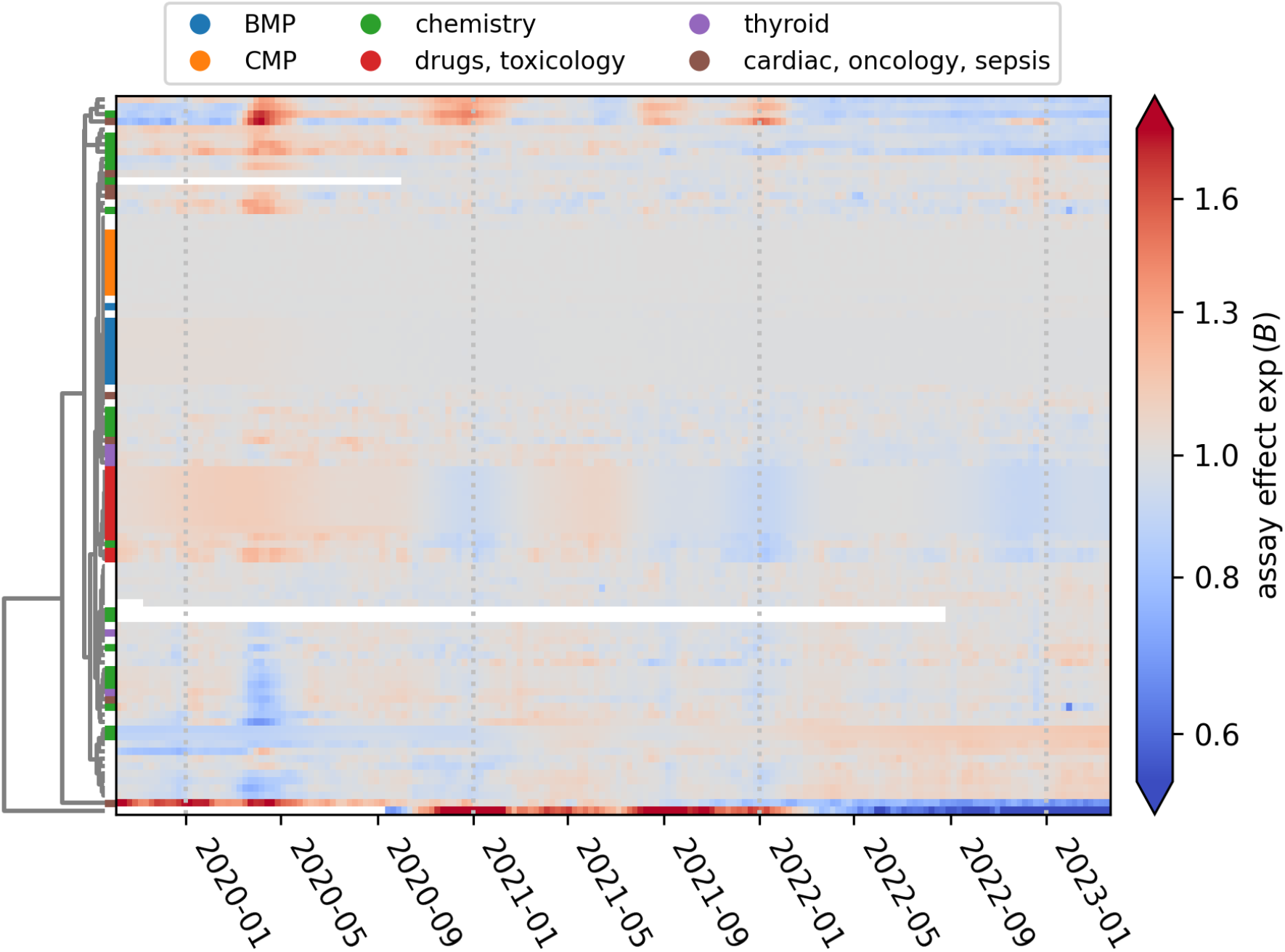
Hierarchical clustering of temporal effects reveals assay groups. Each row of the heat map corresponds to one of 97 different assays and represents the assay-specific effects exp(B). Missing values indicate that the corresponding assay had no observations prior to the corresponding date. Rows are ordered by hierarchical clustering as indicated by the dendrogram on the left, and different assay types are indicated by categorical colors. Assays that belong to the basic metabolic panel (BMP), comprehensive metabolic panel (CMP), and drug and toxicology panel cluster together, reflecting ordering patterns.

To assess the potential for using operational diagnostics data to monitor public health, we identified assays whose temporal effects *B* would have been indicative of SARS-CoV-2 infections during the COVID-19 pandemic. Specifically, we assessed covariation between assay-specific effects and national weekly hospital admissions associated with COVID-19. This data was collated and published by the U.S. Centers for Disease Control and Prevention (CDC) after the first wave of the pandemic in 2020 (U.S. Centers for Disease Control and Prevention, 2024). During the first COVID-19 wave and while non-pharmaceutical interventions, such as social distancing, were in place, many assay volumes were severely depressed in early 2020, as indicated by the common temporal effect *z*_*t*_ shown in panel (a) of Figure 2.

We considered the two assays most correlated and the two assays most anti-correlated with hospital admissions in more detail. The temporal effects *B* for assays used in the triage of respiratory failure, such as procalcitonin for sepsis and C-reactive protein for inflammation, rose during the first and subsequent COVID-19 waves, as shown in panel (b) of Figure 2. The posterior medians of *B* for C-reactive protein and procalcitonin are highly correlated with hospital admissions (Pearson correlation of 0.90 and 0.87, respectively). The volume of follicle-stimulating hormone (FSH) for reproductive endocrinology and lipase assays for pancreatic disease monitoring also covary—although in the opposite direction (Pearson correlation of -0.76 and - 0.77, respectively). Their use decreased even further than the common temporal variation *z*_*t*_. All *p*-values for correlations are < 10^-6^ after Bonferroni correction for multiple hypothesis tests.

We observe similar distinctions between routine and critical care assays during major holidays. For example, the assay-specific holiday effect *ψ* for C-reactive protein and procalcitonin during the week containing Christmas day are 0.036 (95% CI 0.017–0.0554) and 0.087 (95% CI 0.053– 0.121), respectively. Controlling for all other effects, this corresponds to an assay volume increase of 3.6% and 9.1%, respectively. In contrast, for FSH, the posterior median of *ψ* is -0.033 (95% CI - 0.046–-0.019), corresponding to a 3.3% decrease. The posterior median for lipase was 0.099 (95% CI 0.085–0.113), corresponding to a 10.4% increase after controlling for other factors. This observation is consistent with an increase in acute pancreatitis over Christmas as high as 48% (Roberts, et al. 2013).

Our analysis demonstrates that epidemiological signals can be extracted from laboratory assay volume data despite this data not being collected for surveillance purposes. The hierarchical model successfully decomposed assay volumes into interpretable components that reflect the state of public health, as evidenced by the correlation between specific assay patterns and COVID-19 hospitalization data.

This approach offers several advantages over traditional surveillance systems. Data collection incurs no additional costs as it leverages existing infrastructure. There is no additional burden on the monitored population, addressing a key limitation of active surveillance methods. The data structure is consistent across instruments, eliminating the need to harmonize reporting across different healthcare providers, and allowing for data fusion. The data are digital and real-time, reflecting contemporaneous health system responses to evolving epidemiological shifts. Privacy concerns are minimized by focusing solely on aggregate assay volumes rather than individual-level data or test results.

Importantly, however, our system captures only individuals with healthcare access, potentially missing vulnerable populations without healthcare coverage or those with coverage but low levels of health-literacy who fail to access services. The data represent only one vendor’s instruments, and market share may vary demographically or geographically in ways that affect representativeness. Integration of data from other vendors would increase volume and improve population representation. Future work should focus on geographic disaggregation, as national-level changes like those observed during COVID-19 are rare.

While our analysis identified signals correlated with national COVID-19 waves, our framework needs to be validated against other known public health threats, such as seasonal influenza, to assess its utility for broader surveillance. Combining this approach with established monitoring systems could provide complementary signals. Our current analysis cannot assess if operational laboratory diagnostics data can serve as a leading indicator for public health emergencies and epidemiological shifts because the model was fit to the whole dataset. This question that should be addressed in future work through rigorous albeit computationally intensive leave-future-out cross-validation. Development of formal data governance frameworks will be essential to transition this framework from a retrospective analysis to a live, ongoing surveillance platform to augment rather than replace traditional surveillance methods.

## Data Availability

The data supporting the conclusions in this paper is commercially sensitive and was made available to researchers at Harvard University under Non-disclosure Agreement. The code underlying the analyses, including an application to publicly available infectious disease data from Project Tycho, can be found at https://github.com/onnela-lab/sentinel.

## Acknowledgments

All authors were funded in whole or in part by Siemens Healthcare Diagnostics, Inc. (Siemens Healthineers-Harvard Chan RCA, 8317147-01).

## Data and Code Availability

The data supporting the conclusions in this paper is commercially sensitive and was made available to researchers at Harvard University under Non-disclosure Agreement.The code underlying the analyses, including an application to publicly available infectious disease data from Project Tycho, can be found at https://github.com/onnela-lab/sentinel.

## Methods

### Data and Preprocessing

We obtained daily records of assay runs for 1.250 billion assays aggregated by assay type and instrument between 2019-10-07 and 2023-03-26 after removing quality control, specimen validity, and experimental tests. We iteratively processed the dataset further to make it more amenable to analysis.

Assay codes in the dataset were not standardized, and we discarded 0.05% of runs whose assay code did not match a reference table of valid codes. We further dropped low-volume assays that, cumulatively, accounted for less than 1% of runs. For each assay-instrument pair, we removed records prior to the first active week, defined as the first week with any records on five or more days, to mitigate the effect of experimental runs during the setup of new instruments. Records for instrument-assay pairs were removed if they did not have at least ten records, retaining 97.5% of the original dataset. We further removed instruments that did not run at least ten different assays (97.0% retained). For each instrument-assay pair, we filled in missing records between the first and last observation with zeros. Due to desynchronization between Atellica instruments and the backend after the transition to daylight saving time (DST), there were gaps in the data from some instruments until they were restarted. Because the mechanism leading to missing data (change to DST) is not related to the underlying processes, the data are missing completely at random. Consequently, it was possible to discard the impacted records without affecting the analysis (Gelman, Carlin, et al. 2013, ch. 8). For each year, we removed records for each instrument starting in the week of the DST transition up to and including the first week in which any data was recorded again.

### Model Specification and Fitting

The log-linear model for assay volumes in eq. (1) decomposes the three-dimensional tensor *y*_*ijt*_, representing the number of assays performed by instrument *i* of assay *j* in week *t*, into interpretable components, such as common temporal variability *z* shared across all instruments and assays or specialization of instrument *i* to assay *j* in the interaction term *C*_*ij*_. We used a Bayesian approach because it allowed us to incorporate domain knowledge in the form of priors. For example, the common temporal effect *z* in weeks *t* and *t* + 1 are related, and we used a Gaussian process prior to capture the temporal smoothness (Rasmussen and Williams 2006). The model is not identifiable because additive degeneracies, e.g., adding a constant to μ and subtracting the same constant from *a*, leaves predictions unchanged. Adding priors for all parameters breaks the degeneracies and allows the model to be fit. We found sum-to-zero priors (Ogle and Barber 2020) did not aid convergence because the number of observations associated with different instruments and assays varied by orders of magnitude. Priors and descriptions for all parameters are shown in table 1. Log-transformed assay volumes log(1 + *y*) and holiday indicators *X* were standardized prior to model fitting, i.e., transformed to have zero mean and unit variance across all observations. All results are reported on the original, un-standardized scale.

**Table 1:**
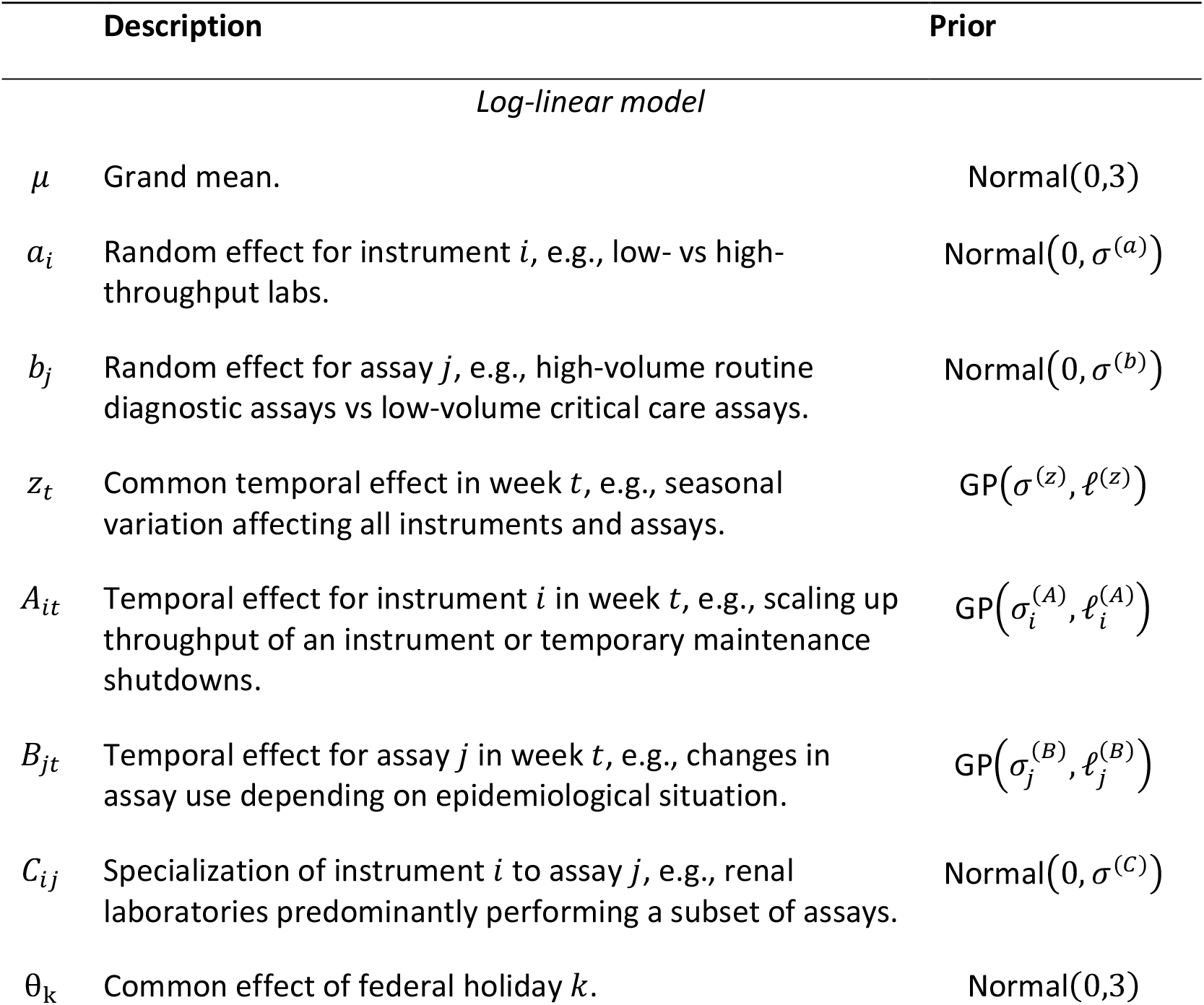
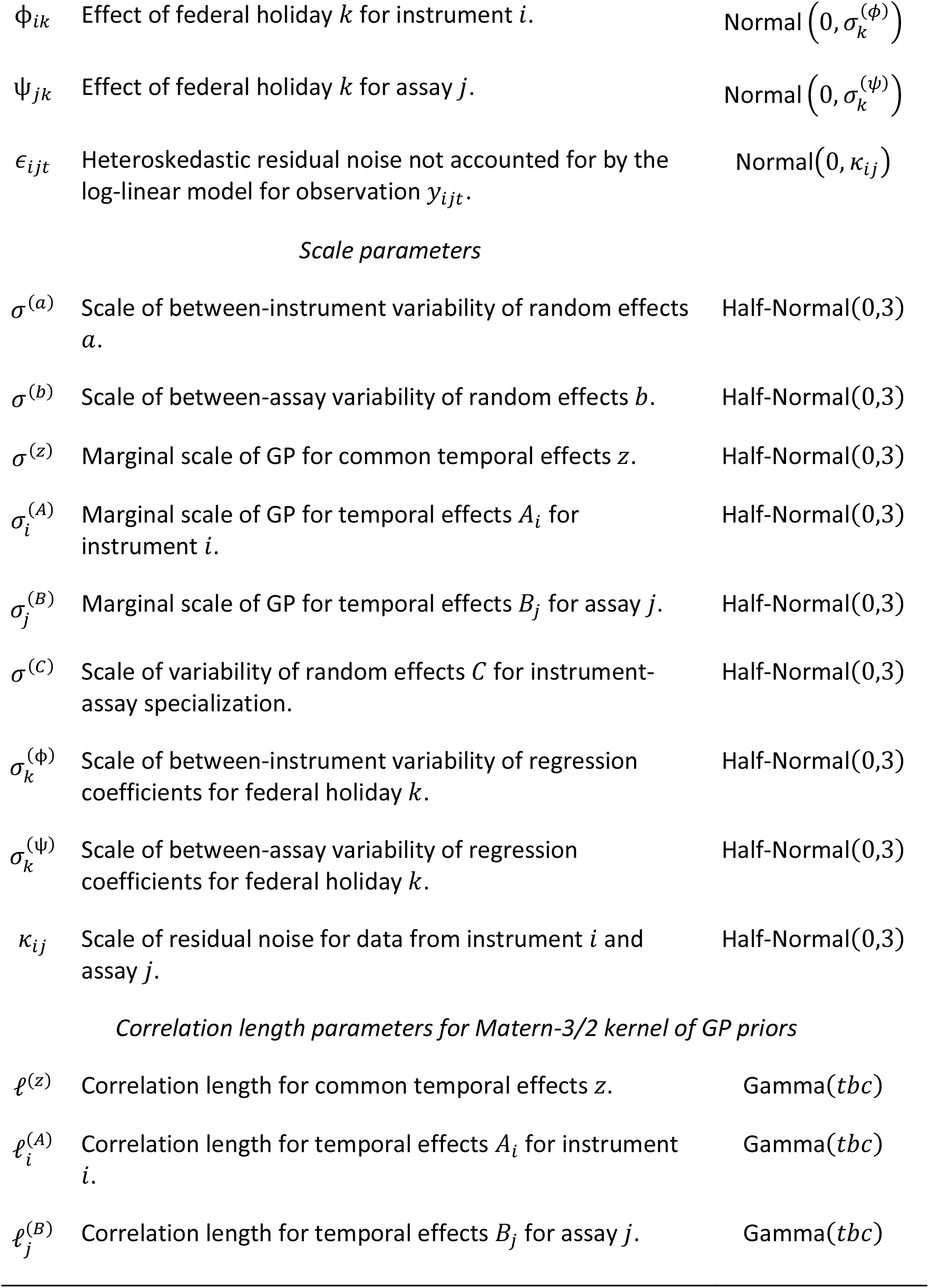
Overview of priors for different components of the model.

| Description |  | Prior |
| --- | --- | --- |
| *Log-linear model* |  |  |
| $\mu$ | Grand mean. | $\text{Normal}(0, 3)$ |
| $a_i$ | Random effect for instrument $i$, e.g., low- vs high-throughput labs. | $\text{Normal}(0, \sigma^{(a)})$ |
| $b_j$ | Random effect for assay $j$, e.g., high-volume routine diagnostic assays vs low-volume critical care assays. | $\text{Normal}(0, \sigma^{(b)})$ |
| $z_t$ | Common temporal effect in week $t$, e.g., seasonal variation affecting all instruments and assays. | $\text{GP}(\sigma^{(z)}, \ell^{(z)})$ |
| $A_{it}$ | Temporal effect for instrument $i$ in week $t$, e.g., scaling up throughput of an instrument or temporary maintenance shutdowns. | $\text{GP}(\sigma_i^{(A)}, \ell_i^{(A)})$ |
| $B_{jt}$ | Temporal effect for assay $j$ in week $t$, e.g., changes in assay use depending on epidemiological situation. | $\text{GP}(\sigma_j^{(B)}, \ell_j^{(B)})$ |
| $C_{ij}$ | Specialization of instrument $i$ to assay $j$, e.g., renal laboratories predominantly performing a subset of assays. | $\text{Normal}(0, \sigma^{(C)})$ |
| $\theta_k$ | Common effect of federal holiday $k$. | $\text{Normal}(0, 3)$ |
| $\phi_{ik}$ | Effect of federal holiday $k$ for instrument $i$. | Normal($0, \sigma_k^{(\phi)}$) |
| $\psi_{jk}$ | Effect of federal holiday $k$ for assay $j$. | Normal($0, \sigma_k^{(\psi)}$) |
| $\epsilon_{ijt}$ | Heteroskedastic residual noise not accounted for by the log-linear model for observation $y_{ijt}$. | Normal($0, \kappa_{ij}$) |
| *Scale parameters* |  |  |
| $\sigma^{(a)}$ | Scale of between-instrument variability of random effects $a$. | Half-Normal(0,3) |
| $\sigma^{(b)}$ | Scale of between-assay variability of random effects $b$. | Half-Normal(0,3) |
| $\sigma^{(z)}$ | Marginal scale of GP for common temporal effects $z$. | Half-Normal(0,3) |
| $\sigma_i^{(A)}$ | Marginal scale of GP for temporal effects $A_i$ for instrument $i$. | Half-Normal(0,3) |
| $\sigma_j^{(B)}$ | Marginal scale of GP for temporal effects $B_j$ for assay $j$. | Half-Normal(0,3) |
| $\sigma^{(C)}$ | Scale of variability of random effects $C$ for instrument-assay specialization. | Half-Normal(0,3) |
| $\sigma_k^{(\phi)}$ | Scale of between-instrument variability of regression coefficients for federal holiday $k$. | Half-Normal(0,3) |
| $\sigma_k^{(\psi)}$ | Scale of between-assay variability of regression coefficients for federal holiday $k$. | Half-Normal(0,3) |
| $\kappa_{ij}$ | Scale of residual noise for data from instrument $i$ and assay $j$. | Half-Normal(0,3) |
| *Correlation length parameters for Matern-3/2 kernel of GP priors* |  |  |
| $\ell^{(z)}$ | Correlation length for common temporal effects $z$. | Gamma($tbc$) |
| $\ell_i^{(A)}$ | Correlation length for temporal effects $A_i$ for instrument $i$. | Gamma($tbc$) |
| $\ell_j^{(B)}$ | Correlation length for temporal effects $B_j$ for assay $j$. | Gamma($tbc$) |

The posterior distribution is not tractable, and we used black-box variational inference to approximate it (Ranganath, Gerrish and Blei 2014). In short, the posterior is approximated by a normal distribution in an unconstrained space, and the posterior approximation is obtained by transforming random variables to the correct domain. For example, the observation noise variance κ is approximated by a normal distribution in log-space, such that the posterior approximation is a log-normal distribution. The posterior approximation was initialized as a mean-field normal distribution with location parameter sampled from a uniform distribution on the interval [−0.01,0.01] and initial scale of 0.001. The model was implemented in Python using the NumPyro probabilistic programming language (Phan, Pradhan and Jankowiak 2019). We used the Adam optimizer (Kingma and Ba 2014) to minimize the Kullback-Leibler divergence between the true and approximate posterior on the full dataset because minibatch training gave rise to sparse gradients, hampering convergence. The optimization was run for 10 million iterations, starting with a learning rate of 10^−2^. The learning rate was reduced to 10^−3^ and 10^−4^ at 7 million and 9 million iterations, respectively.

We took two steps to accelerate fitting the model. First, we collapsed the intercept *μ*^;^ and common temporal effects *z* to reduce the number of parameters. Recall that *μ* ∼Normal (0, *σ* ^(*μ;*)^) and *z*∼MVN (0, *K*^(*z*)^), where MVN (0, *K*^(*z*)^) denotes the multivariate normal distribution with covariance matrix *K*^(*z*)^. We use a Matérn-3/2 kernel for the Gaussian process such that the covariance of *z* at times *t* and *t*^′^ 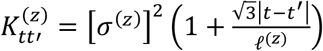 exp 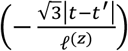, where |*t* − *t*^’^ | denotes the absolute distance between *t* and *t*^’^ in weeks. Then the collapsed variable *ζ* = *μ; + z*_*t*_ has zero mean and covariance 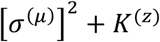. We similarly collapsed *a* with *A* and *b* with *B*. Second, we approximated the Gaussian process likelihood using a kernel with circular boundary conditions which facilitates efficient evaluation of the likelihood using Fourier methods (Hoffmann and Onnela 2025). To attenuate the effect of periodic boundary conditions, we padded the temporal domain from 181 weeks by a third to 242 weeks.

### Assay Clustering

We clustered assays based on the similarity of their temporal effects *B* using a hierarchical clustering algorithm with complete linkage implemented in SciPy (Virtanen, et al. 2020). The distance measure between assays *j* and *k* was the mean absolute difference of temporal effects

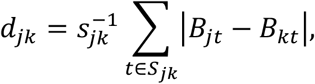

where *S*_jk_ is the set of weeks where a signal for both assays is available, and *s*_jk_ is the size of the set. We consider a signal to be available after the first week in which any runs of the assay were recorded after the preprocessing described above.

## Appendices

### Table of *R*^2^ for different assays

The table shows the correlation coefficient between exp(*B*) and weekly national COVID-19 hospital admissions. The coefficient of determination *R*^2^ is based on a linear regression model predicting hospital admissions given exp(*B*). While the correlation coefficient may be negative,*R*^2^ is always positive because even exp(*B*) that are anti-correlated with the admission data can explain variance in the latter.

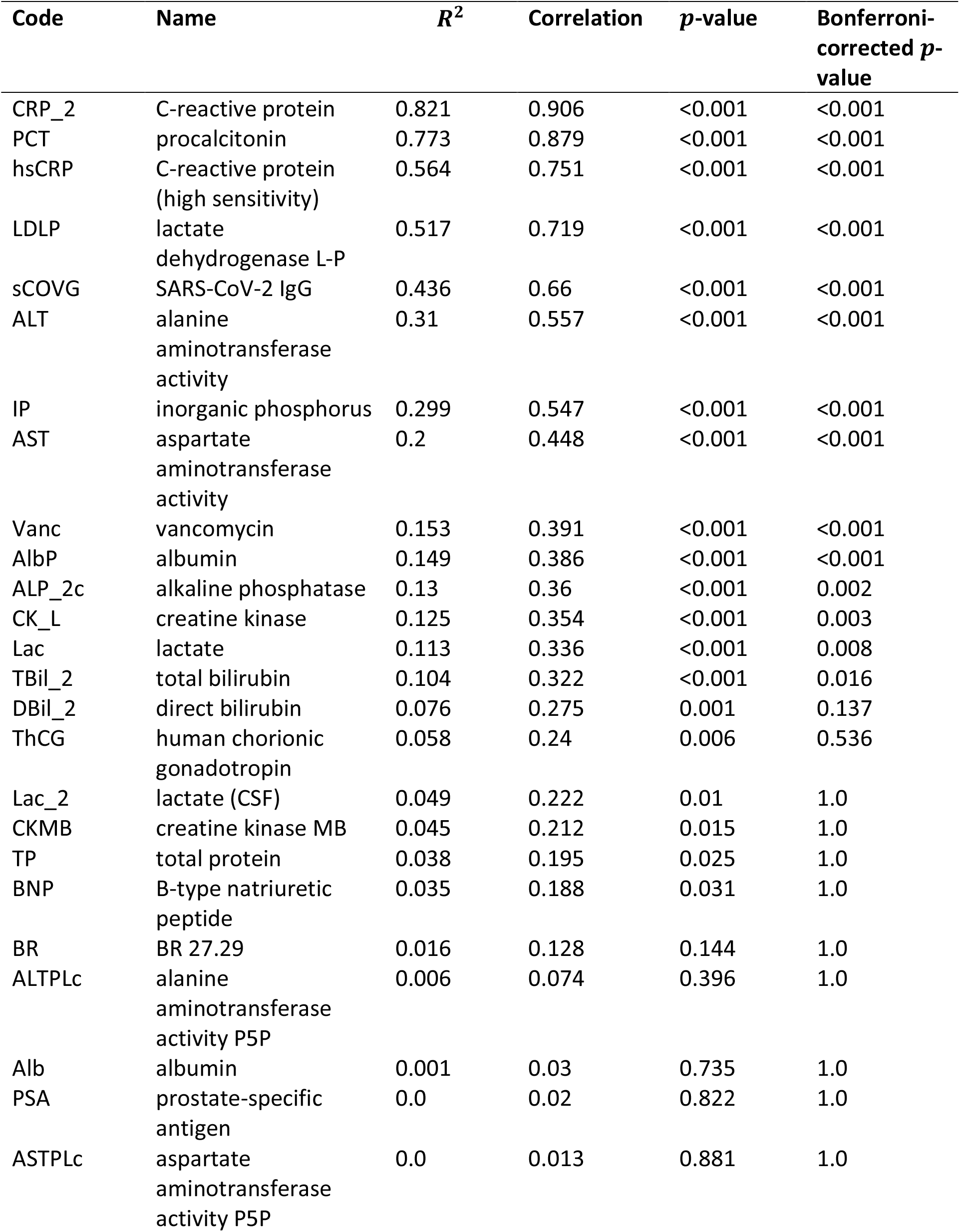
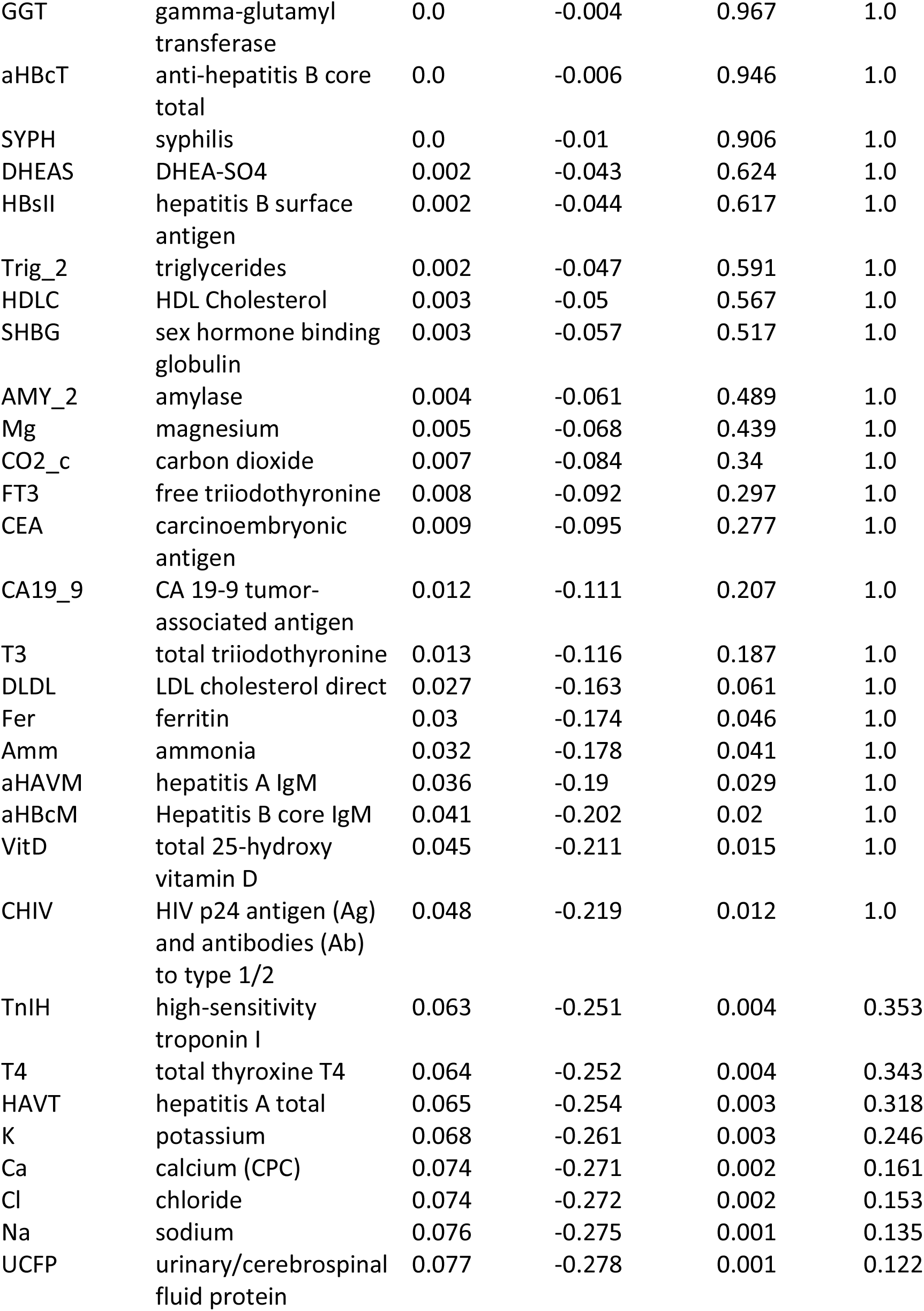
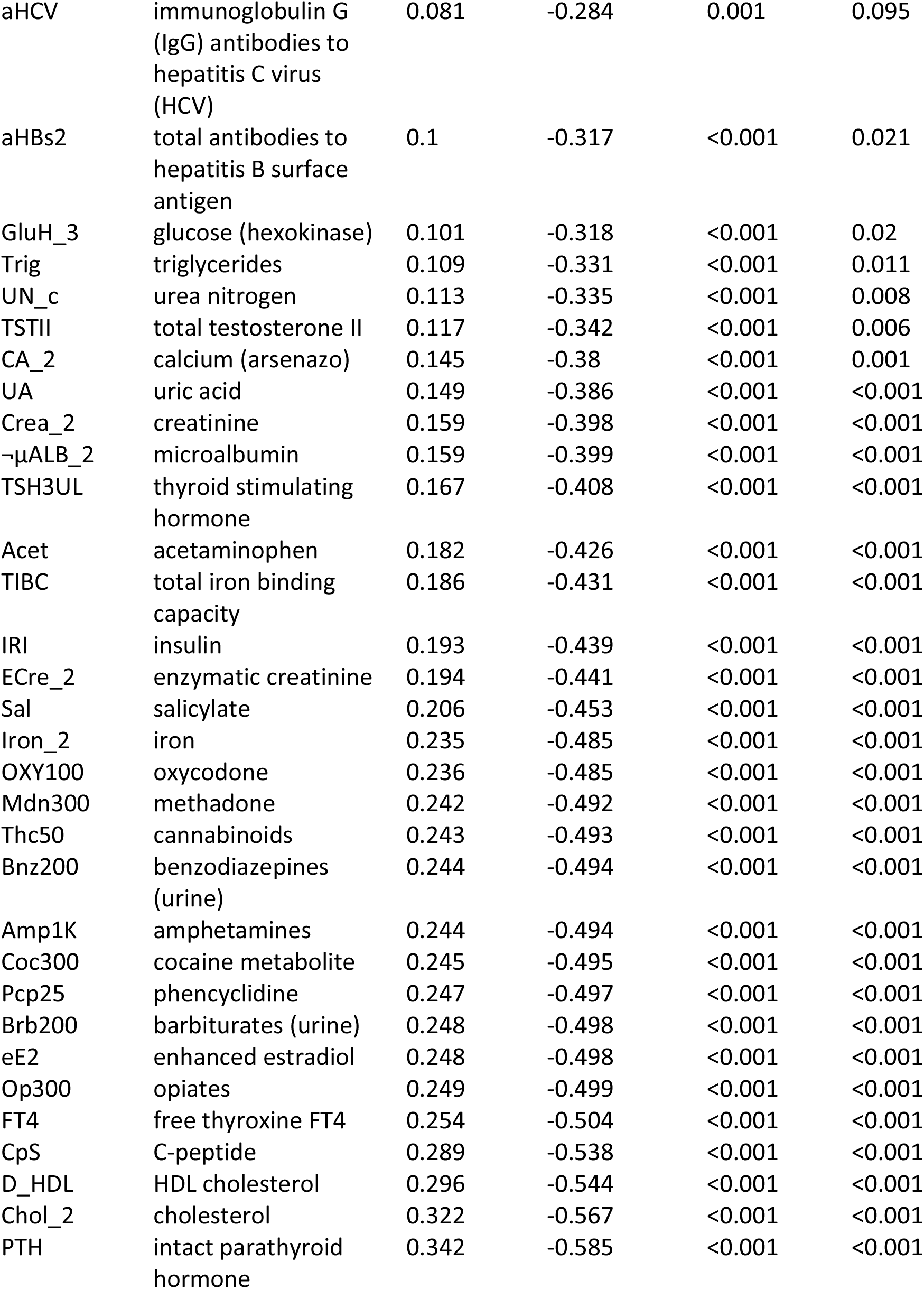
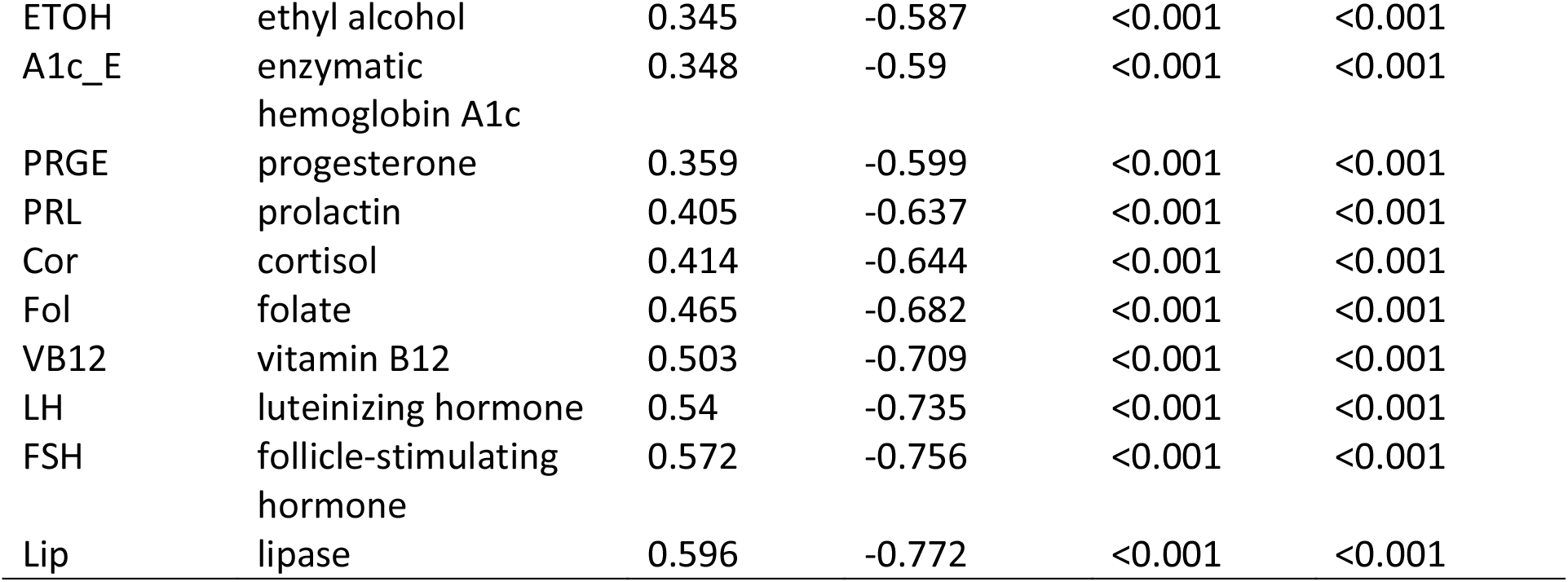

| Code | Name | $R^2$ | Correlation | $p$-value | Bonferroni-corrected $p$-value |
| --- | --- | --- | --- | --- | --- |
| CRP_2 | C-reactive protein | 0.821 | 0.906 | <0.001 | <0.001 |
| PCT | procalcitonin | 0.773 | 0.879 | <0.001 | <0.001 |
| hsCRP | C-reactive protein (high sensitivity) | 0.564 | 0.751 | <0.001 | <0.001 |
| LDLP | lactate dehydrogenase L-P | 0.517 | 0.719 | <0.001 | <0.001 |
| sCOVG | SARS-CoV-2 IgG | 0.436 | 0.66 | <0.001 | <0.001 |
| ALT | alanine aminotransferase activity | 0.31 | 0.557 | <0.001 | <0.001 |
| IP | inorganic phosphorus | 0.299 | 0.547 | <0.001 | <0.001 |
| AST | aspartate aminotransferase activity | 0.2 | 0.448 | <0.001 | <0.001 |
| Vanc | vancomycin | 0.153 | 0.391 | <0.001 | <0.001 |
| AlbP | albumin | 0.149 | 0.386 | <0.001 | <0.001 |
| ALP_2c | alkaline phosphatase | 0.13 | 0.36 | <0.001 | 0.002 |
| CK_L | creatine kinase | 0.125 | 0.354 | <0.001 | 0.003 |
| Lac | lactate | 0.113 | 0.336 | <0.001 | 0.008 |
| TBil_2 | total bilirubin | 0.104 | 0.322 | <0.001 | 0.016 |
| DBil_2 | direct bilirubin | 0.076 | 0.275 | 0.001 | 0.137 |
| ThCG | human chorionic gonadotropin | 0.058 | 0.24 | 0.006 | 0.536 |
| Lac_2 | lactate (CSF) | 0.049 | 0.222 | 0.01 | 1.0 |
| CKMB | creatine kinase MB | 0.045 | 0.212 | 0.015 | 1.0 |
| TP | total protein | 0.038 | 0.195 | 0.025 | 1.0 |
| BNP | B-type natriuretic peptide | 0.035 | 0.188 | 0.031 | 1.0 |
| BR | BR 27.29 | 0.016 | 0.128 | 0.144 | 1.0 |
| ALTPLc | alanine aminotransferase activity P5P | 0.006 | 0.074 | 0.396 | 1.0 |
| Alb | albumin | 0.001 | 0.03 | 0.735 | 1.0 |
| PSA | prostate-specific antigen | 0.0 | 0.02 | 0.822 | 1.0 |
| ASTPLc | aspartate aminotransferase activity P5P | 0.0 | 0.013 | 0.881 | 1.0 |
| GGT | gamma-glutamyl transferase | 0.0 | -0.004 | 0.967 | 1.0 |
| aHBcT | anti-hepatitis B core total | 0.0 | -0.006 | 0.946 | 1.0 |
| SYPH | syphilis | 0.0 | -0.01 | 0.906 | 1.0 |
| DHEAS | DHEA-SO4 | 0.002 | -0.043 | 0.624 | 1.0 |
| HBsII | hepatitis B surface antigen | 0.002 | -0.044 | 0.617 | 1.0 |
| Trig_2 | triglycerides | 0.002 | -0.047 | 0.591 | 1.0 |
| HDLC | HDL Cholesterol | 0.003 | -0.05 | 0.567 | 1.0 |
| SHBG | sex hormone binding globulin | 0.003 | -0.057 | 0.517 | 1.0 |
| AMY_2 | amylase | 0.004 | -0.061 | 0.489 | 1.0 |
| Mg | magnesium | 0.005 | -0.068 | 0.439 | 1.0 |
| CO2_c | carbon dioxide | 0.007 | -0.084 | 0.34 | 1.0 |
| FT3 | free triiodothyronine | 0.008 | -0.092 | 0.297 | 1.0 |
| CEA | carcinoembryonic antigen | 0.009 | -0.095 | 0.277 | 1.0 |
| CA19_9 | CA 19-9 tumor-associated antigen | 0.012 | -0.111 | 0.207 | 1.0 |
| T3 | total triiodothyronine | 0.013 | -0.116 | 0.187 | 1.0 |
| DLDL | LDL cholesterol direct | 0.027 | -0.163 | 0.061 | 1.0 |
| Fer | ferritin | 0.03 | -0.174 | 0.046 | 1.0 |
| Amm | ammonia | 0.032 | -0.178 | 0.041 | 1.0 |
| aHAVM | hepatitis A IgM | 0.036 | -0.19 | 0.029 | 1.0 |
| aHBcM | Hepatitis B core IgM | 0.041 | -0.202 | 0.02 | 1.0 |
| VitD | total 25-hydroxy vitamin D | 0.045 | -0.211 | 0.015 | 1.0 |
| CHIV | HIV p24 antigen (Ag) and antibodies (Ab) to type 1/2 | 0.048 | -0.219 | 0.012 | 1.0 |
| TnIH | high-sensitivity troponin I | 0.063 | -0.251 | 0.004 | 0.353 |
| T4 | total thyroxine T4 | 0.064 | -0.252 | 0.004 | 0.343 |
| HAVT | hepatitis A total | 0.065 | -0.254 | 0.003 | 0.318 |
| K | potassium | 0.068 | -0.261 | 0.003 | 0.246 |
| Ca | calcium (CPC) | 0.074 | -0.271 | 0.002 | 0.161 |
| Cl | chloride | 0.074 | -0.272 | 0.002 | 0.153 |
| Na | sodium | 0.076 | -0.275 | 0.001 | 0.135 |
| UCFP | urinary/cerebrospinal fluid protein | 0.077 | -0.278 | 0.001 | 0.122 |
| aHCV | immunoglobulin G (IgG) antibodies to hepatitis C virus (HCV) | 0.081 | -0.284 | 0.001 | 0.095 |
| aHBs2 | total antibodies to hepatitis B surface antigen | 0.1 | -0.317 | <0.001 | 0.021 |
| GluH_3 | glucose (hexokinase) | 0.101 | -0.318 | <0.001 | 0.02 |
| Trig | triglycerides | 0.109 | -0.331 | <0.001 | 0.011 |
| UN_c | urea nitrogen | 0.113 | -0.335 | <0.001 | 0.008 |
| TSTII | total testosterone II | 0.117 | -0.342 | <0.001 | 0.006 |
| CA_2 | calcium (arsenazo) | 0.145 | -0.38 | <0.001 | 0.001 |
| UA | uric acid | 0.149 | -0.386 | <0.001 | <0.001 |
| Crea_2 | creatinine | 0.159 | -0.398 | <0.001 | <0.001 |
| -μALB_2 | microalbumin | 0.159 | -0.399 | <0.001 | <0.001 |
| TSH3UL | thyroid stimulating hormone | 0.167 | -0.408 | <0.001 | <0.001 |
| Acet | acetaminophen | 0.182 | -0.426 | <0.001 | <0.001 |
| TIBC | total iron binding capacity | 0.186 | -0.431 | <0.001 | <0.001 |
| IRI | insulin | 0.193 | -0.439 | <0.001 | <0.001 |
| ECre_2 | enzymatic creatinine | 0.194 | -0.441 | <0.001 | <0.001 |
| Sal | salicylate | 0.206 | -0.453 | <0.001 | <0.001 |
| Iron_2 | iron | 0.235 | -0.485 | <0.001 | <0.001 |
| OXY100 | oxycodone | 0.236 | -0.485 | <0.001 | <0.001 |
| Mdn300 | methadone | 0.242 | -0.492 | <0.001 | <0.001 |
| Thc50 | cannabinoids | 0.243 | -0.493 | <0.001 | <0.001 |
| Bnz200 | benzodiazepines (urine) | 0.244 | -0.494 | <0.001 | <0.001 |
| Amp1K | amphetamines | 0.244 | -0.494 | <0.001 | <0.001 |
| Coc300 | cocaine metabolite | 0.245 | -0.495 | <0.001 | <0.001 |
| Pcp25 | phencyclidine | 0.247 | -0.497 | <0.001 | <0.001 |
| Brb200 | barbiturates (urine) | 0.248 | -0.498 | <0.001 | <0.001 |
| eE2 | enhanced estradiol | 0.248 | -0.498 | <0.001 | <0.001 |
| Op300 | opiates | 0.249 | -0.499 | <0.001 | <0.001 |
| FT4 | free thyroxine FT4 | 0.254 | -0.504 | <0.001 | <0.001 |
| CpS | C-peptide | 0.289 | -0.538 | <0.001 | <0.001 |
| D_HDL | HDL cholesterol | 0.296 | -0.544 | <0.001 | <0.001 |
| Chol_2 | cholesterol | 0.322 | -0.567 | <0.001 | <0.001 |
| PTH | intact parathyroid hormone | 0.342 | -0.585 | <0.001 | <0.001 |
| ETOH | ethyl alcohol | 0.345 | -0.587 | <0.001 | <0.001 |
| A1c_E | enzymatic<br>hemoglobin A1c | 0.348 | -0.59 | <0.001 | <0.001 |
| PRGE | progesterone | 0.359 | -0.599 | <0.001 | <0.001 |
| PRL | prolactin | 0.405 | -0.637 | <0.001 | <0.001 |
| Cor | cortisol | 0.414 | -0.644 | <0.001 | <0.001 |
| Fol | folate | 0.465 | -0.682 | <0.001 | <0.001 |
| VB12 | vitamin B12 | 0.503 | -0.709 | <0.001 | <0.001 |
| LH | luteinizing hormone | 0.54 | -0.735 | <0.001 | <0.001 |
| FSH | follicle-stimulating<br>hormone | 0.572 | -0.756 | <0.001 | <0.001 |
| Lip | lipase | 0.596 | -0.772 | <0.001 | <0.001 |

